# Revolutionizing COPD and Asthma Management with Artificial Intelligence

**DOI:** 10.1101/2025.03.18.25324219

**Authors:** Yudi Kurniawan Budi Susilo, Shamima Abdul Rahman

## Abstract

The integration of artificial intelligence (AI) into the management of chronic obstructive pulmonary disease (COPD) and asthma offers significant advancements in patient care, diagnosis, and treatment personalization. AI technologies, particularly machine learning and deep learning, have shown great promise in predictive modeling, enabling earlier detection and more accurate diagnoses. AI-driven tools, such as telemedicine and remote monitoring applications, are helping clinicians provide personalized care and manage these conditions more effectively, especially for high-risk patients. Additionally, AI’s ability to analyze vast datasets, including electronic health records and medical imaging, allows for more dynamic and adaptive treatment plans tailored to individual patients’ needs. However, challenges remain, such as ensuring data privacy, addressing ethical concerns, and developing standardized regulatory frameworks to support the implementation of AI in clinical settings. Despite these challenges, the potential of AI to revolutionize COPD and asthma management is vast, and continued research and collaboration will be critical in overcoming these barriers and enhancing healthcare outcomes for patients worldwide.

## 1 Introduction

The application of artificial intelligence (AI) in the management of chronic respiratory diseases, particularly chronic obstructive pulmonary disease (COPD) and asthma, is rapidly evolving and holds significant promise for improving patient outcomes. AI technologies are increasingly being integrated into diagnostic processes, treatment personalization, and remote monitoring capabilities, ultimately transforming the approach to managing these chronic conditions.

AI systems have demonstrated considerable advancements in predictive modeling for asthma and COPD. Kothalawala et al. highlighted a machine learning-based Childhood Asthma Prediction Program (CAPP) exhibiting a 30% improvement in sensitivity over traditional models, indicating stronger predictive capabilities for pediatric asthma management (Kothalawala et al., 2021). The systematic review by Kothalawala et al. also emphasizes the importance of leveraging diverse predictors within machine learning frameworks, which can uncover novel biomarkers and improve model robustness (Kothalawala et al., 2020). Moreover, the accuracy of disease classification has been significantly enhanced through the integration of electronic health record (EHR) data into machine learning models, as highlighted by Liu et al. (Liu et al., 2024).

Telemedicine and remote monitoring applications represent another frontier for AI integration in managing COPD and asthma. Graña-Castro et al. identified that AI-driven tools can facilitate early diagnosis and empower personalized treatment strategies, critical for enhancing health-related quality of life and reducing hospitalizations through timely interventions (Graña-Castro et al., 2024). In line with these findings, Alanazi et al. provide insight into how AI algorithms can inform treatment plans tailored to patient-specific data, emphasizing the transformative potential of AI in real-time patient care and management (Alanazi et al., 2024). Furthermore, while Villafuerte et al. discuss the application of AI and IoT technologies in monitoring and diagnosing respiratory infections, this highlights their interdisciplinary nature in modern healthcare solutions (Villafuerte et al., 2023).

In addition to optimizing treatment delivery, AI plays a significant role in understanding disease progression. Machine learning models utilizing vital sign trajectories have been shown to predict respiratory failure and assist with decision-making in critical care settings, thereby improving patient outcomes during emergencies (Kim et al., 2019). These predictive analytics are also echoed in Stivi et al.’s investigation, which discusses the potential of AI in predicting weaning success from mechanical ventilation, underscoring its relevance in acute care scenarios (Stivi et al., 2024).

Ethical considerations and data management challenges persist as AI technologies become more prevalent in healthcare. The integration of AI must address issues related to data quality, patient privacy, and the need for robust regulatory frameworks to ensure the safe use of these technologies (Graña-Castro et al., 2024). Therefore, while the potential of AI in chronic respiratory disease management is significant, ongoing research and dialogue around these ethical implications are crucial for responsible implementation.

In conclusion, the integration of AI into the management of COPD and asthma is not just enhancing clinical decision-making and treatment personalization, but is also paving the way for innovative approaches to patient monitoring and disease prediction. The efficacy of AI-dependent strategies reflects a transformative shift in the healthcare landscape that prioritizes data-driven, patient-centric care.

## 2 Literature

The integration of artificial intelligence (AI) and machine learning (ML) in the management of chronic obstructive pulmonary disease (COPD) and asthma has rapidly emerged as a transformative approach, facilitating enhanced diagnostics, personalized treatment plans, and improved patient outcomes. This revolution is largely driven by the vast amounts of data generated through medical records, respiratory sound analysis, and imaging technologies, as well as advancements in computational techniques. It is essential to explore how current research leverages AI to address the complexities of these respiratory diseases, focusing on diagnostic accuracy, predictive modeling, and the prospect of personalized medicine.

Early studies have demonstrated the efficacy of machine learning algorithms in diagnosing asthma and COPD with substantial specificity and sensitivity. For instance, Spathis and Vlamos reported that the Random Forest classifier achieved a diagnostic precision of 80.3% for asthma, identifying key factors such as maximum expiratory flow, smoking status, age, and wheezing as significant attributes influencing diagnosis (Spathis & Vlamos, 2017). The findings were supported by other studies highlighting the effectiveness of Random Forest in accurately detecting asthma compared to conventional methods (Spathis & Vlamos, 2017). Furthermore, advancements in deep learning have generated improved diagnostic frameworks, enhancing traditional ML approaches by employing models such as Long Short-Term Memory (LSTM) networks, which can analyze temporal data patterns to predict disease outcomes more effectively (Yu et al., 2021).

The utilization of electronic health records (EHRs) to train machine learning models presents a significant opportunity for predicting the recurrence of respiratory pathologies, enriching current clinical practices with data-driven insights (Rodríguez et al., 2021). Chronic lower respiratory diseases often exhibit heterogeneous characteristics necessitating precision in management. Unsupervised machine learning techniques have successfully identified latent clusters among patients with asthma and COPD, enabling clinicians to implement targeted interventions based on identifiable phenotypes (Cui et al., 2020). This tailoring of treatment, complemented by continuous data collection through devices like wearables and the Internet of Medical Things (IoMT), allows for dynamic adjustments in therapeutic strategies as patient conditions evolve (Santana-Mancilla et al., 2023).

Moreover, digital health strategies have shown promise in improving asthma monitoring through AI-enhanced platforms that analyze patient data for more accurate control assessments (Novembre et al., 2022). By systematically tracking symptoms and medication adherence, these technologies empower both patients and healthcare providers to engage in more informed dialogues about disease management, thereby facilitating proactive interventions. This is especially relevant in low- and middle-income countries (LMICs), where access to updated management strategies for asthma is often limited (Ozoh et al., 2022). By employing AI, healthcare systems can adapt to local needs, improving the overall management framework for respiratory diseases.

The intersection of AI and medical imaging represents another frontier for enhancing COPD and asthma management. Deep neural networks (DNNs) have been effectively applied to analyze chest radiographs and other imaging data, facilitating the identification of pathological conditions that might otherwise go unnoticed (Gonem et al., 2020). Studies illustrate that such models can achieve diagnostic capabilities exceeding traditional image analysis methods, providing a basis for timely and accurate assessments of patients’ respiratory health (Bansal, 2021).

Additionally, acoustic signal analysis has emerged as a novel avenue for classifying respiratory disorders, harnessing ML algorithms to analyze respiratory sounds such as wheezing, coughing, and other abnormal patterns. Techniques like convolutional neural networks (CNNs) have demonstrated high effectiveness in differentiating between various lung conditions based on audio inputs, presenting opportunities for non-invasive diagnostics (Sfayyih et al., 2023;, Srivastava et al., 2021). The ability to detect respiratory anomalies through sound can significantly enhance early intervention efforts, ultimately reducing morbidity associated with respiratory diseases.

Another pivotal aspect revolves around the predictive capabilities of AI in assessing risks associated with respiratory diseases, particularly in older adults. Machine learning models validated on respiratory frequency data have illustrated profound potential for forecasting respiratory events, which is crucial for preemptive healthcare measures (Santana-Mancilla et al., 2023). These predictive analytics enable healthcare providers to develop tailored care pathways, ensuring that at-risk populations receive timely interventions personalized to their specific health profiles.

The development and implementation of tailored predictive models for respiratory diseases necessitate rigorous training algorithms capable of managing complex datasets. Advances in methods such as Support Vector Machines (SVM) and other supervised learning frameworks have proven instrumental in constructing reliable prediction systems that categorize patients based on risk assessment models (Uddin et al., 2019). Notably, the implementation of these systems has been correlated with enhanced clinical decision-making, ensuring that patients with severe cases of COPD or asthma can receive immediate and appropriate care (Li et al., 2022).

Each of these advancements underscores the critical importance of a multidisciplinary approach in integrating AI into respiratory care. Collaborative efforts between computer scientists, healthcare professionals, and regulatory bodies are crucial in navigating the ethical and practical considerations inherent in utilizing AI in medicine. As the landscape of COPD and asthma management continues to evolve, ongoing research efforts are pivotal in validating the effectiveness of these technologies while simultaneously addressing barriers to their adoption in clinical settings.

The future of COPD and asthma management lies in effectively harnessing the vast potential of AI to enhance diagnostic accuracy and treatment efficacy while transforming patient engagement through personalized care pathways powered by real-time data analytics. AI-driven interventions have the power to streamline healthcare resources while improving disease management outcomes, thereby revolutionizing the way respiratory diseases are addressed across diverse healthcare settings.

In conclusion, the integration of AI and machine learning into the management of COPD and asthma represents a key milestone in the evolution of respiratory medicine. While challenges remain in terms of implementation, validation, and ethical considerations, the promising advancements documented in the literature indicate that with continued research and collaborative efforts, significant improvements in respiratory health outcomes can be achieved.

## 3 Method

The methodology outlines a systematic approach to conducting a bibliometric analysis focused on the application of artificial intelligence (AI) in the management, treatment, diagnosis, monitoring, and prediction of chronic obstructive pulmonary disease (COPD) and asthma. The analysis is based on literature retrieved from the Scopus database, with a specific focus on journal articles published in English between 2021 and 2025. The search strategy employed the following keywords: “COPD,” “chronic obstructive pulmonary disease,” “asthma,” “artificial intelligence,” “AI,” “machine learning,” “deep learning,” “management,” “treatment,” “diagnosis,” “monitoring,” and “prediction.”

**Figure 1:**
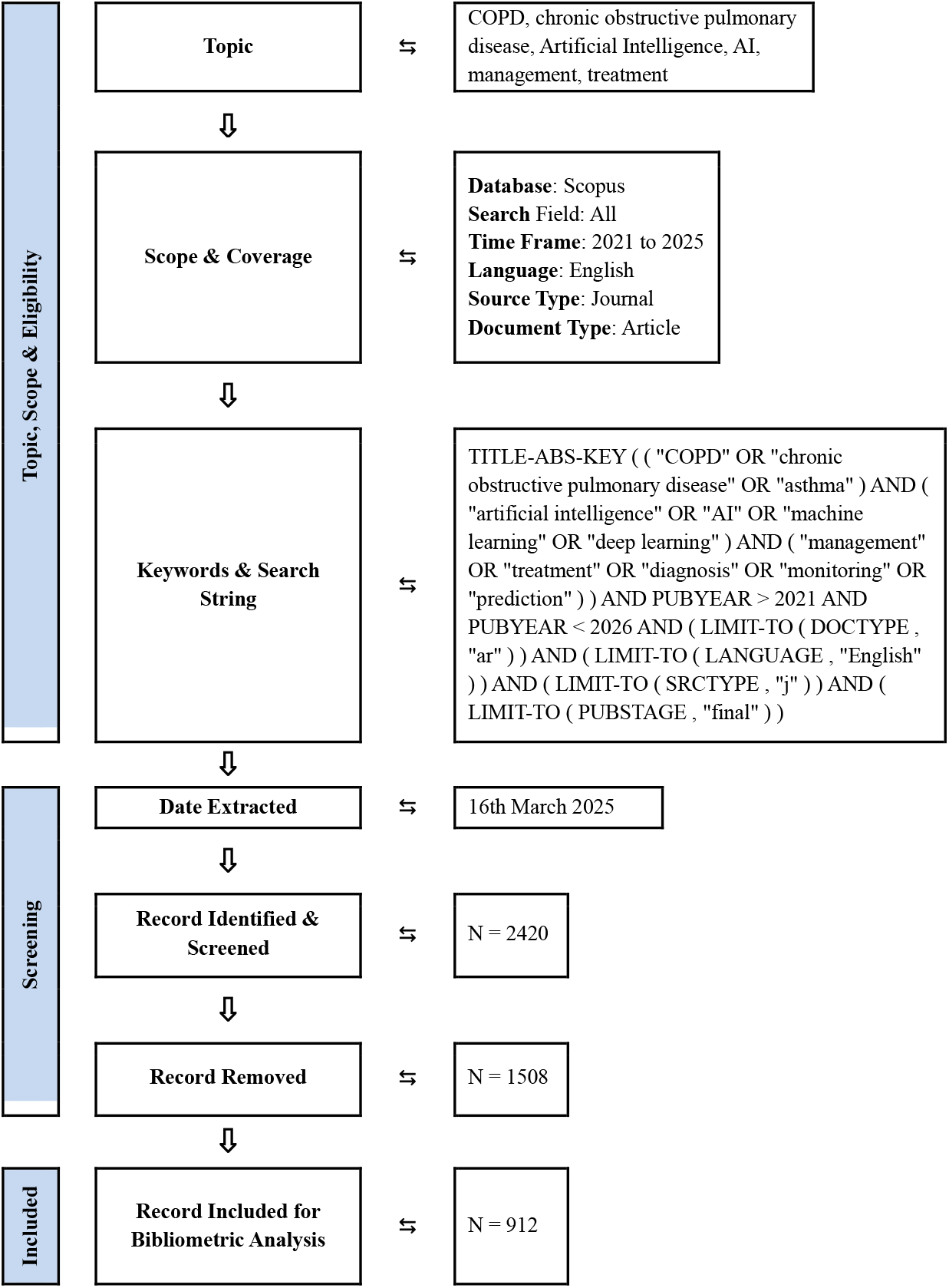
Flow diagram of the search strategy for keyword “COPD”, “chronic obstructive pulmonary disease”, “Artificial Intelligence”, “AI”, “management, treatment”

The initial search yielded 2,420 records, which were subsequently screened. After removing 1,508 records that did not meet the inclusion criteria, 912 records were included for bibliometric analysis. The inclusion criteria were based on relevance to the specified topics, document type (articles), language (English), and publication stage (final). The exclusion criteria removed records that did not align with these parameters.

The analysis utilized specialized software tools such as VOSviewer, Harzing’s Publish or Perish and Bibliometrix (R) to conduct citation analysis, keyword co-occurrence analysis, and author collaboration network mapping. These tools enabled the visualization of citation networks and the identification of significant research trends, providing insights into the development of AI applications in COPD and asthma management.

Potential biases and limitations were acknowledged, including the reliance on Scopus as the sole database, which may affect the generalizability of the findings. The methodology adheres to established best practices, with appropriate citation of the PRISMA framework and relevant bibliometric studies, ensuring methodological transparency and rigor. The date of data extraction was March 16, 2025.

## 4 Result

The provided document results, reveal a dynamic publication trend over the past four years. Starting with 216 documents in 2022, the publication output steadily increased to 265 in 2023, peaking at 349 in 2024. However, a significant drop is observed in 2025, with only 82 documents recorded, likely due to the data extraction occurring early in the year (March 16, 2025), not capturing the full annual output. This trend suggests a growing interest and research activity in the application of artificial intelligence to manage chronic respiratory diseases, with a notable surge in publications leading up to 2024.

**Figure 2:**
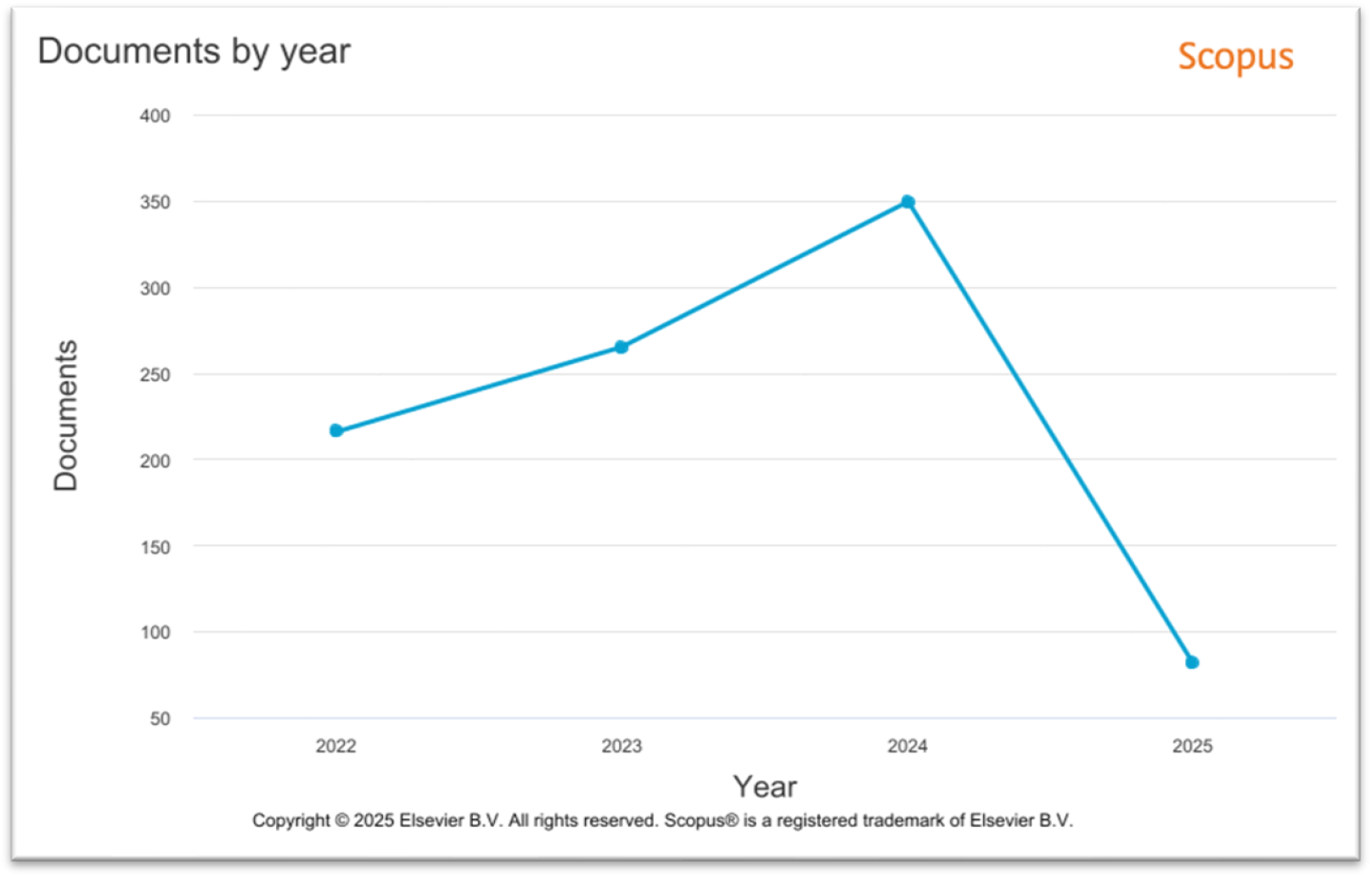
Document dynamic publication trend over the past four years (2022 to 2025)

The fluctuation in document numbers highlights the evolving nature of research in this field and the potential impact of emerging trends and technologies on publication output. The analysis provides a comprehensive overview of citation networks, keyword co-occurrences, and author collaborations, offering valuable insights into the development of AI applications in COPD and asthma management. However, it’s important to acknowledge the limitations of relying solely on the Scopus database, which might not fully represent the global research landscape.

### 4.1 Document by subject area

A significant portion, 38.1%, falls under the Medicine category, highlighting the core focus of this research on medical applications. Computer Science represents the second largest segment at 13.3%, indicating the substantial role of computational methods and AI in this field. Engineering follows with 9.3%, suggesting a focus on developing technological solutions and devices. Biochemistry, Genetics, and Molecular Biology contribute 8.0%, reflecting the biological and molecular aspects of respiratory diseases and their interaction with AI-driven interventions.

**Figure 2:**
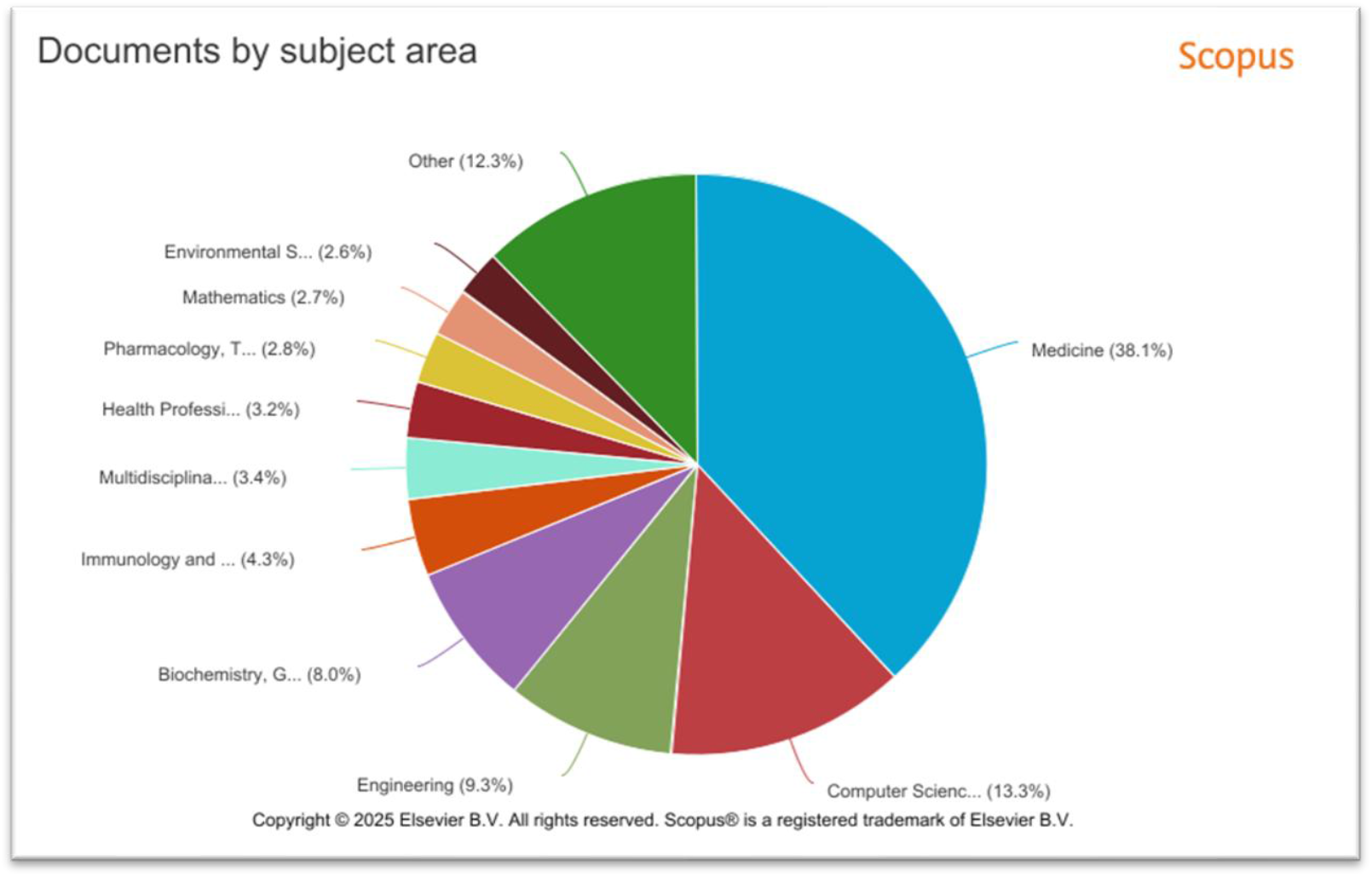
Document by subject area

Immunology and Microbiology comprise 4.3%, emphasizing the immunological factors in COPD and asthma and their relevance to AI-based management strategies. Multidisciplinary research accounts for 3.4%, indicating the interdisciplinary nature of this field. Health Professions contribute 3.2%, highlighting the practical applications of AI in healthcare settings. Pharmacology, Toxicology, and Pharmaceutics represent 2.8%, suggesting the involvement of drug development and delivery in AI-assisted treatments. Mathematics and Environmental Science both account for 2.7% and 2.6% respectively, indicating the quantitative modelling and environmental factors considered in this research.

A substantial portion, 12.3%, is categorized as “Other,” which likely encompasses a variety of smaller, less dominant subject areas that still contribute to the overall research landscape. This distribution underscores the multidisciplinary approach required to advance AI-driven solutions for COPD and asthma management, with Medicine, Computer Science, and Engineering playing pivotal roles.

### 4.2 Author by country

The United States leads significantly in research output with 232 documents, demonstrating a substantial focus on AI applications in COPD and asthma management. This is further supported by their high citation count of 1516 and normalized citation count of 257.6114, indicating a strong impact and recognition of their work within this field.

**Figure 3:**
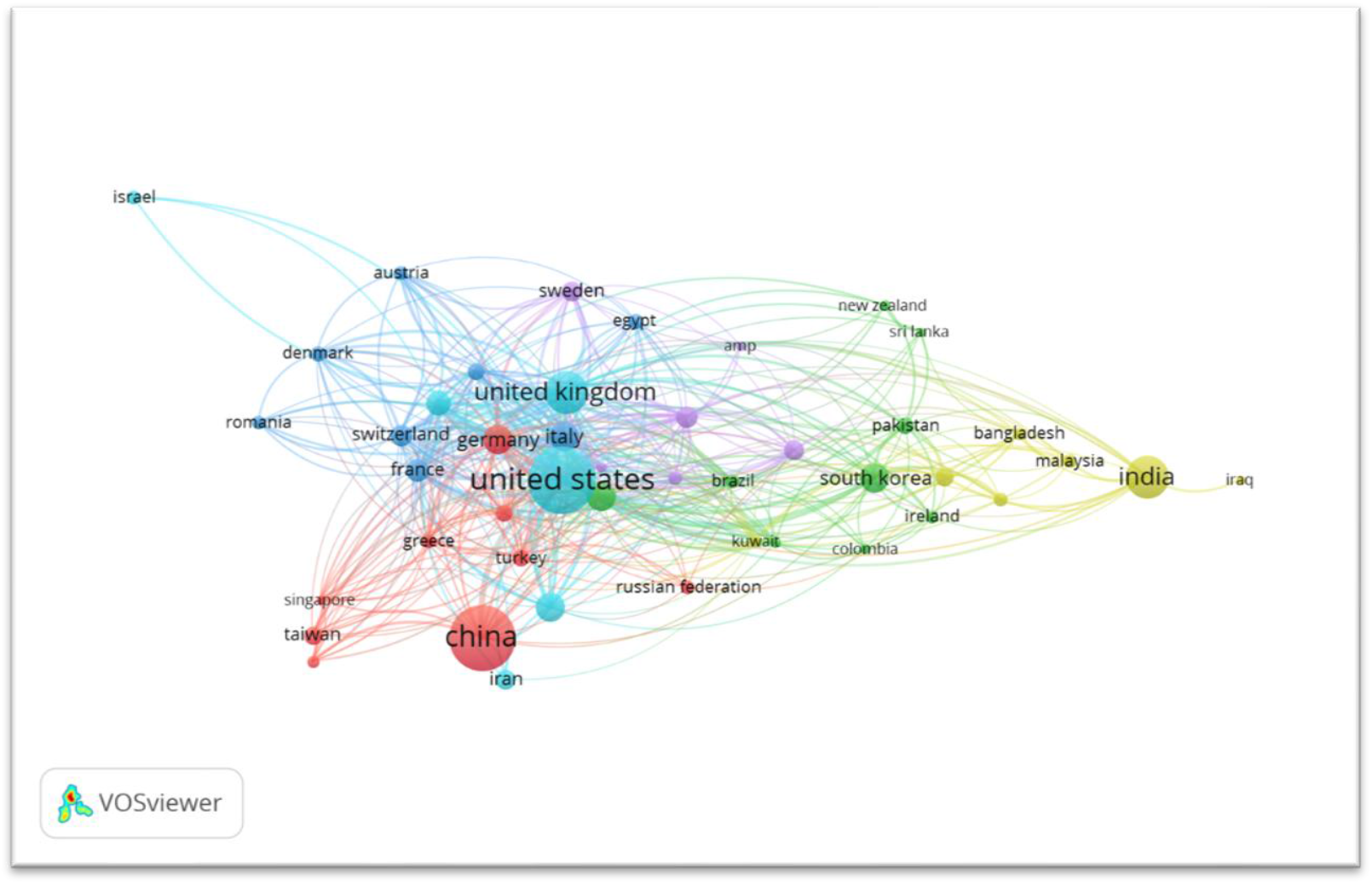
Collaboration among authors and research output of different countries

China follows closely in document count with 216 publications, suggesting a rapidly growing interest and research activity in AI-driven solutions for respiratory diseases. Their citation count of 832 and normalized citation count of 177.2791 reflect a significant contribution to the field, albeit with a slightly lower impact per publication compared to the US.

The United Kingdom shows a substantial contribution with 96 documents, and their citation metrics (673 citations, 122.1278 normalized citations) indicate a high-quality, impactful research output. India also demonstrates a significant research focus with 92 documents, 584 citations, and 104.2718 normalized citations, highlighting its growing role in this domain.

Several European countries, including Italy, Germany, and Spain, contribute noticeably to the research landscape. Italy, with 46 documents and high normalized citations (93.7483), indicates impactful research. Germany and Canada have similar document counts (45 each) but differ in citation impact, with Germany showing a lower normalized citation count (55.2091) compared to Canada (65.5256). Spain stands out with 393 citations and 117.8207 normalized citations from 43 documents, suggesting a high-impact contribution.

South Korea and the Netherlands also contribute, with South Korea showing a moderate impact (48.868 normalized citations) and the Netherlands having a lower but still relevant contribution (32.3527 normalized citations).

Overall, the United States and China are leading in terms of research output, while the US, UK, and Spain demonstrate a higher impact in terms of normalized citations. This analysis underscores the global effort in advancing AI applications for COPD and asthma management, with significant contributions from various countries.

### 4.3 Bibliographic coupling by document among authors

The analysis offers a clear view of the influence and interconnectedness of the authors contributing to the field of COPD and asthma management using artificial intelligence (AI). This analysis, based on citation counts, highlights both established leaders and emerging contributors in this evolving research area.

**Figure 4:**
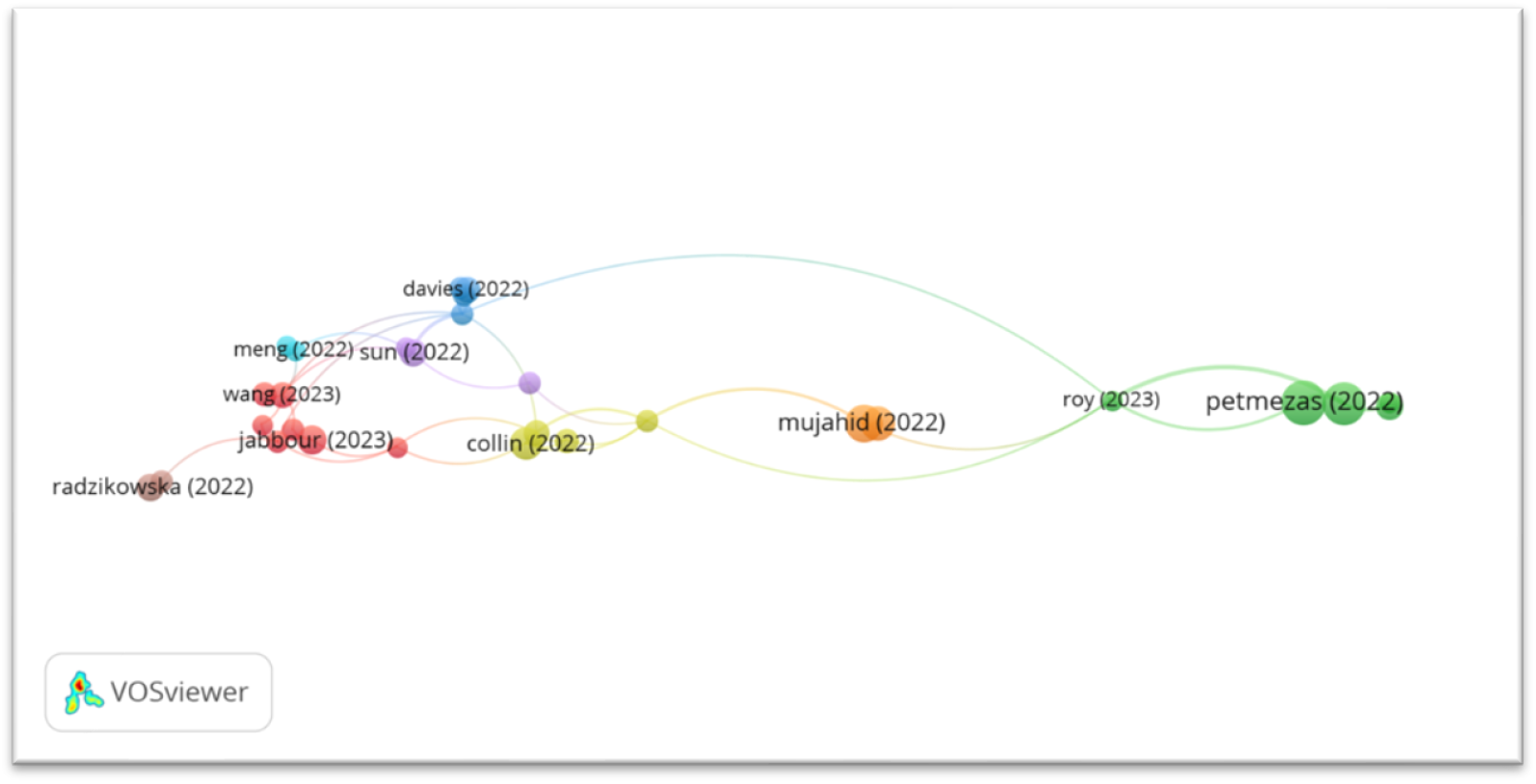
Network visualization map of the bibliographic coupling between authors

The author with the highest citation counts, such as Petmezas (2022) (98 citations) and Nguyen (2022) (91 citations), represent the cornerstone of AI research in the management of COPD and asthma. These works are likely to have laid the foundation for key AI applications in the field, whether through the development of predictive models, machine learning algorithms, or other AI-driven solutions that enhance clinical decision-making and patient care. Their research, often referenced by others, plays a critical role in shaping the methodologies and technologies currently being utilized in AI-assisted respiratory disease management.

On the other hand, researchers like Mujahid (2022) (76 citations), Bhosale (2023) (58 citations), and Collin (2022) (53 citations) have contributed significantly, though their works are cited less frequently. This suggests that while their research may be more recent or specialized, it still plays an important role in expanding the scope of AI applications in the field. As AI technologies in healthcare continue to evolve, their contributions may gain greater recognition and influence, particularly in areas of innovative algorithms or specific therapeutic techniques in COPD and asthma management.

Meanwhile, authors with lower citation counts, such as Jabour (2023), Radzikowski (2022), Dash (2022), Sun (2022), and Shamji (2023), may be contributing research that is more niche or in the early stages of becoming recognized in the broader academic community. While their works are not yet as widely cited, they represent the ongoing progression of the field and may offer new perspectives, methods, or applications that are essential for the future development of AI in respiratory disease management.

This analysis serves as a valuable tool to understand the current landscape of research and the interconnectedness of authors within the domain of AI and respiratory disease management. It not only showcases the influential works that have driven the field forward but also points to the emerging voices whose contributions will likely play an increasingly vital role in the future of AI-driven healthcare solutions. As the body of research continues to expand, the interplay between established and emerging researchers will help catalyze further advancements, ultimately revolutionizing the management of COPD and asthma through innovative AI technologies.

### 4.4 Authors keyword

The keyword analysis reveals a strong emphasis on machine learning and artificial intelligence (AI) in the research on COPD and asthma management. The term machine learning stands out with 290 occurrences, underscoring its pivotal role in advancing AI technologies for healthcare applications, particularly in predicting patient outcomes, personalizing treatment strategies, and analyzing complex medical data. Asthma follows closely with 125 occurrences, indicating that a significant portion of research is dedicated to asthma management, where AI can enhance diagnostic accuracy, monitoring, and treatment optimization.

**Figure 4:**
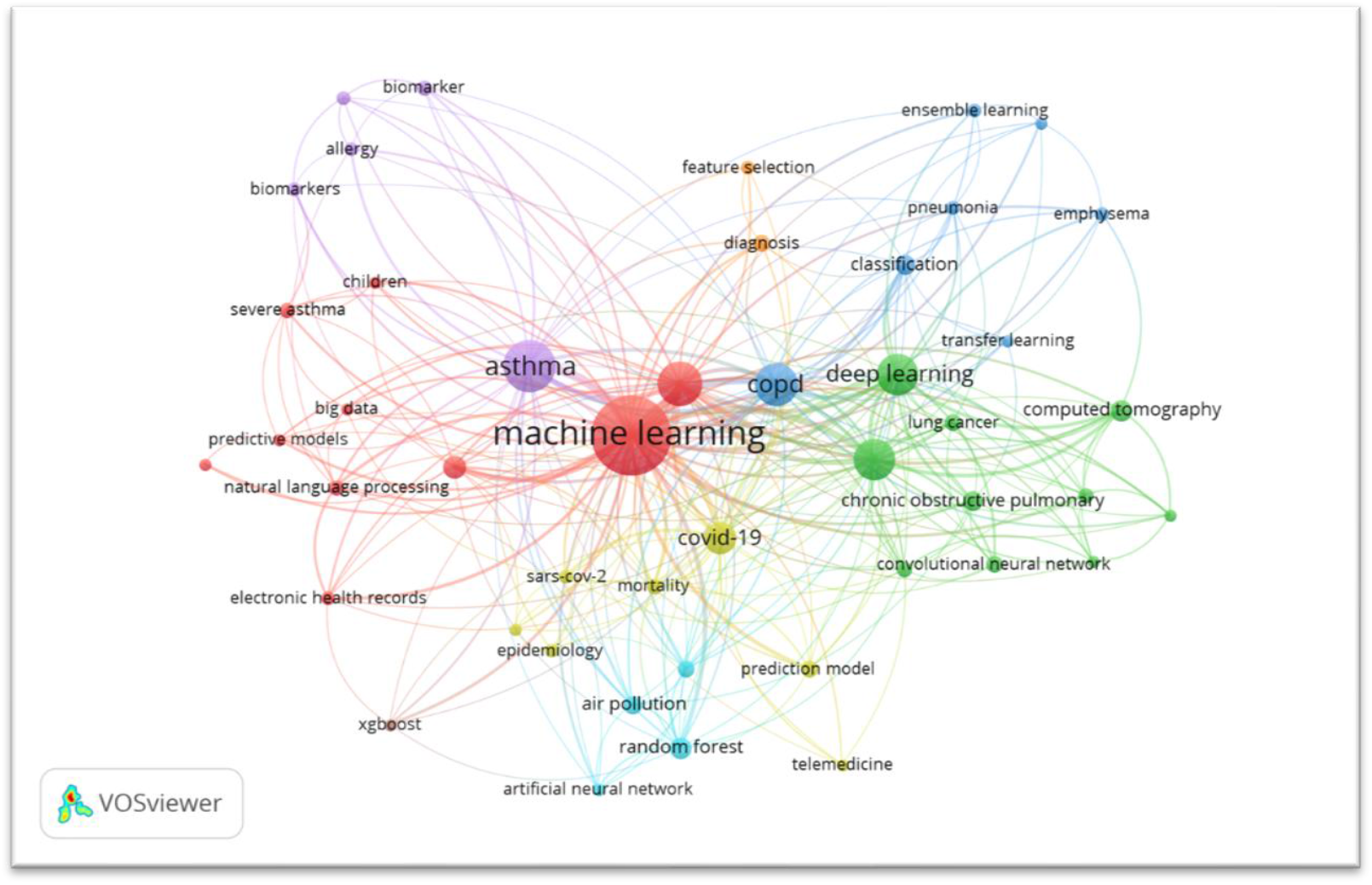
Network visualization map of the bibliographic coupling of authors keywords

The term artificial intelligence itself appears 91 times, reflecting the broad application of AI techniques across the field, including machine learning and deep learning, which are central to improving healthcare outcomes. COPD, with 86 occurrences, highlights the importance of AI in managing chronic obstructive pulmonary disease, a major respiratory condition that can benefit from AI-driven solutions such as early detection and predictive modeling. A slightly more specific term, chronic obstructive pulmonary disease, appears 79 times, further emphasizing the focus on this condition within the research landscape.

Deep learning, with 79 occurrences, suggests that more advanced AI methods are being applied in tasks like medical imaging and complex data analysis, critical for improving the diagnosis and management of respiratory diseases. The presence of COVID-19 among the keywords (48 occurrences) points to the significant impact of the pandemic on respiratory research, with AI being explored for predicting outcomes, managing patients, and analyzing lung scans associated with the virus.

Prediction is also an important theme, mentioned 26 times, highlighting the growing interest in AI-based predictive models for forecasting disease progression and patient-specific treatment responses. Computed tomography, appearing 21 times, reflects the use of advanced medical imaging technologies in combination with AI to analyze lung conditions and detect abnormalities. Finally, random forest, also appearing 21 times, indicates that machine learning algorithms, such as random forests, are being employed to build predictive models for disease classification and risk assessment.

Overall, the keyword analysis illustrates that AI, particularly through machine learning and deep learning, is central to research efforts aimed at improving the management of COPD and asthma. The frequent mention of specific technologies like computed tomography and algorithms like random forests highlights the technical depth of current research, while the inclusion of COVID-19 shows how recent global health challenges have influenced the focus of this field.

### 4.5 Highly cited articles

The table highlights several highly cited articles that demonstrate the growing role of artificial intelligence (AI) in revolutionizing the management of chronic obstructive pulmonary disease (COPD) and asthma. With the integration of AI technologies, particularly machine learning and deep learning techniques, the healthcare industry is seeing substantial advancements in diagnosing, classifying, and predicting outcomes for patients with respiratory diseases like COPD and asthma.

**Table 1:**
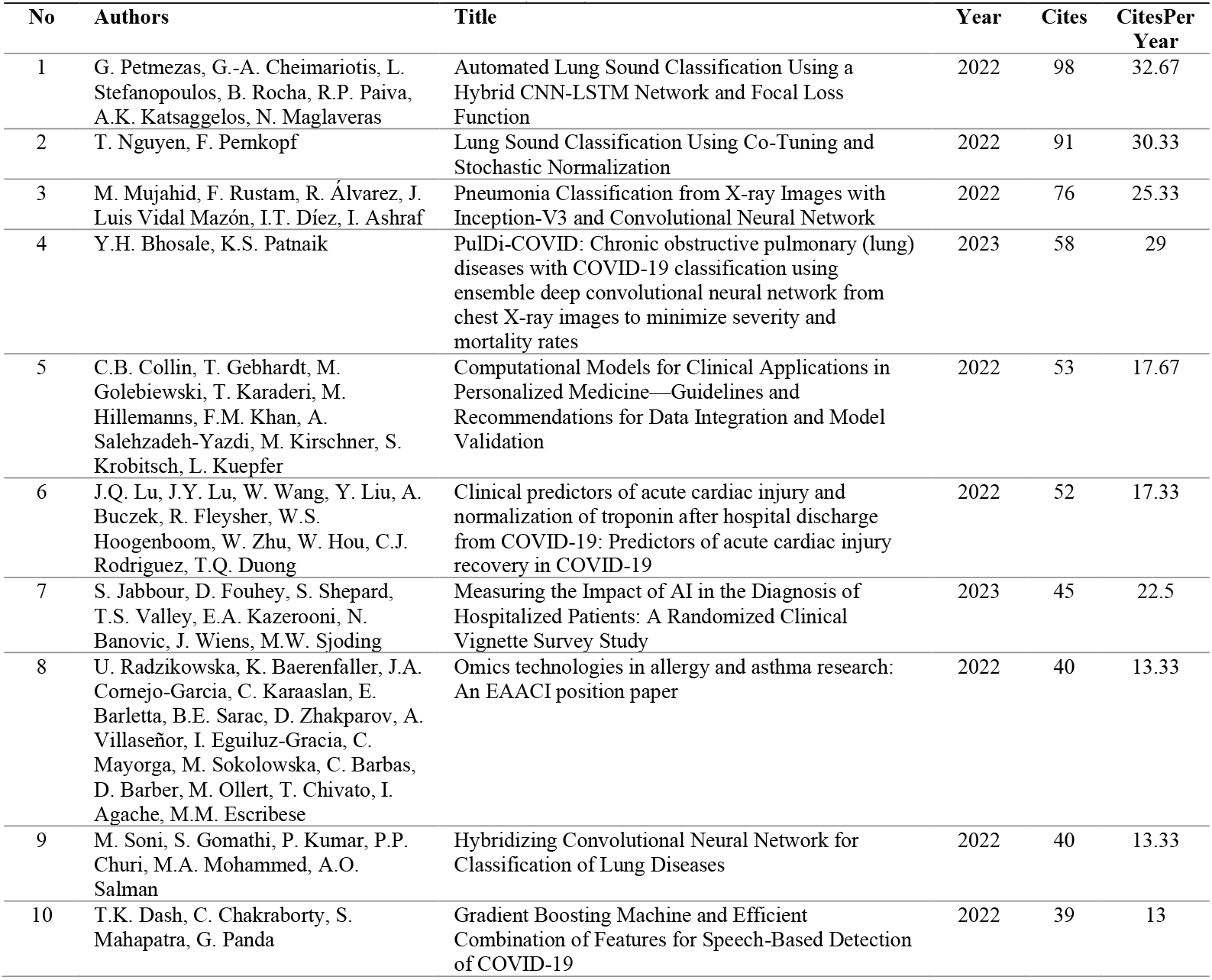
The growing role of artificial intelligence (AI) in revolutionizing the management of chronic obstructive pulmonary disease (COPD) and asthma.

For instance, the article “PulDi-COVID: Chronic obstructive pulmonary (lung) diseases with COVID-19 classification” by Y.H. Bhosale and K.S. Patnaik, published in 2023, shows how AI is being used to classify COPD and other pulmonary diseases in the context of COVID-19.

This research has already garnered 58 citations, reflecting its significance in addressing the complex challenges faced by healthcare providers when managing COPD patients during the pandemic. The article’s citation rate of 29 per year further suggests that it is rapidly gaining traction as a vital tool in the management of respiratory diseases.

Additionally, the research on lung sound classification using AI, such as “Automated Lung Sound Classification Using a Hybrid CNN-LSTM Network and Focal Loss Function” by G. Petmezas et al., is essential for non-invasive diagnosis and real-time monitoring of conditions like asthma and COPD. With 98 citations and a citation rate of 32.67 per year, this study exemplifies the potential of AI in detecting lung conditions early and accurately, allowing for more precise management of asthma and COPD.

Moreover, AI’s application in pneumonia classification, as seen in studies like “Pneumonia Classification from X-ray Images with Inception-V3 and Convolutional Neural Network” by

M. Mujahid et al., underscores the broader implications for AI in managing respiratory diseases by enabling faster and more reliable detection of complications that may arise in COPD and asthma patients. This paper, with 76 citations and a citation rate of 25.33 per year, further supports AI’s growing role in enhancing diagnostic capabilities for respiratory conditions.

These articles collectively highlight the transformative potential of AI in improving the diagnosis, treatment, and management of COPD and asthma, not just through enhancing clinical predictions but also by making healthcare more proactive and personalized. As AI continues to evolve, its integration into COPD and asthma management is expected to lead to better patient outcomes, more efficient treatments, and a deeper understanding of respiratory diseases.

## 5 Discussion

The integration of artificial intelligence (AI) into the management of chronic obstructive pulmonary disease (COPD) and asthma has gained considerable momentum, offering transformative potential for both diagnostic and therapeutic approaches. AI-driven solutions, particularly machine learning (ML) and deep learning (DL), have significantly enhanced diagnostic precision, enabling earlier detection and more personalized treatment options for patients with these chronic respiratory conditions.

One of the key advancements in AI applications is the integration of electronic health records (EHRs) and real-time data from wearable devices to monitor patient conditions continuously. Studies have shown that machine learning models can utilize such data to predict exacerbations, making it possible to intervene proactively and reduce the frequency of hospitalizations. For instance, research by Kothalawala et al. (2021) demonstrated a 30% improvement in sensitivity in predicting pediatric asthma episodes using AI, thus improving the overall management of asthma in children.

The use of AI in analyzing respiratory sounds, such as wheezing and coughing, has also emerged as a non-invasive method to diagnose conditions like asthma and COPD. The work by Petmezas et al. (2022), utilizing a hybrid CNN-LSTM network, exemplifies how AI can identify lung sounds and detect abnormalities associated with these diseases. This approach allows for more accessible and timely diagnostics, particularly in remote areas where healthcare resources may be limited. AI’s ability to classify lung sounds accurately provides a significant opportunity for improving early detection and disease management, making it a critical tool in the future of respiratory care.

Furthermore, the implementation of AI technologies in telemedicine has shown promise in enhancing patient care remotely. AI-powered platforms can now assist in monitoring patients’ symptoms and adherence to prescribed treatments, providing real-time feedback to healthcare providers and patients. This trend is especially beneficial for patients with COPD, as continuous monitoring can help predict exacerbations before they lead to critical conditions. For example, the research by Graña-Castro et al. (2024) highlighted that AI-driven telemedicine tools could empower patients and clinicians to make data-driven decisions that improve health outcomes and reduce hospital visits.

Despite the significant advancements, several challenges remain in the widespread adoption of AI in healthcare. Issues related to data quality, privacy concerns, and the need for robust regulatory frameworks pose significant barriers. AI models require vast amounts of high-quality data for accurate predictions and ensuring patient privacy while utilizing such data remains a major concern. Additionally, the lack of standardized regulatory frameworks for AI applications in healthcare complicates the implementation of these technologies in clinical settings.

Ethical considerations also play a crucial role in the deployment of AI in COPD and asthma management. The need for transparency in AI decision-making processes, as well as the potential for algorithmic bias, must be addressed to ensure equitable access to AI-powered healthcare tools. As AI models are increasingly relied upon for clinical decision-making, it is essential that these technologies are developed and validated through rigorous research to minimize risks and maximize their benefit to patients.

In conclusion, the potential of AI in revolutionizing the management of COPD and asthma is vast. AI-driven approaches are already enhancing diagnostics, predicting disease progression, and personalizing treatment strategies. However, ongoing research and collaboration between healthcare professionals, data scientists, and regulatory bodies are essential to overcoming the challenges of data quality, privacy, and ethics. As these barriers are addressed, AI has the potential to significantly improve the quality of care for patients with COPD and asthma, transforming how these chronic respiratory diseases are managed in the future.

## 6 Conclusion

The integration of artificial intelligence (AI) into the management of chronic obstructive pulmonary disease (COPD) and asthma is reshaping the landscape of respiratory healthcare. AI technologies, particularly machine learning (ML) and deep learning (DL), offer significant advancements in diagnostic accuracy, treatment personalization, and patient monitoring, providing more effective and proactive approaches to managing these chronic conditions. From the use of machine learning algorithms to predict asthma exacerbations to the application of AI in analyzing lung sounds and medical imaging, AI is enhancing both the early detection and management of COPD and asthma.

Telemedicine and remote monitoring powered by AI are particularly promising, enabling continuous, real-time monitoring of patient data. This allows for more timely interventions, potentially reducing hospital admissions and improving overall health outcomes for patients with respiratory diseases. Furthermore, AI’s ability to analyze large datasets, such as electronic health records and respiratory data, enables more tailored treatment plans, leading to better disease management and a more personalized approach to care.

Despite the tremendous potential, there are challenges that must be addressed before AI can be fully integrated into routine clinical practice. Issues such as data privacy, the need for high-quality data, and the lack of standardized regulatory frameworks need to be overcome. Additionally, ethical concerns regarding transparency and potential biases in AI models must be carefully managed to ensure that these technologies are used equitably and responsibly.

In conclusion, AI is poised to revolutionize COPD and asthma management by providing more accurate, timely, and personalized care. While challenges remain, the continuous development of AI technologies and their successful integration into healthcare systems will likely lead to substantial improvements in the quality of life for patients and the efficiency of healthcare delivery. As these technologies continue to evolve, they promise to be a cornerstone of future respiratory disease management, enhancing clinical outcomes and transforming the overall healthcare experience for individuals with COPD and asthma.

## Supporting information

Scopus analyze by subject

Scopus analyze by subject

Scopus analyze by year

Scopus analyze by year

Scopus analyze by source

Bibliographic coupling of documents

Scopus analyze by authors keyword

Scopus analyze by authors country

Scopus dataset

## Data Availability

https://doi.org/10.17632/9z8k2f95f6.1

