## Supplementary figures and images for "Revolutionizing COPD and Asthma Management with Artificial Intelligence"

### Scopus analyze by subject

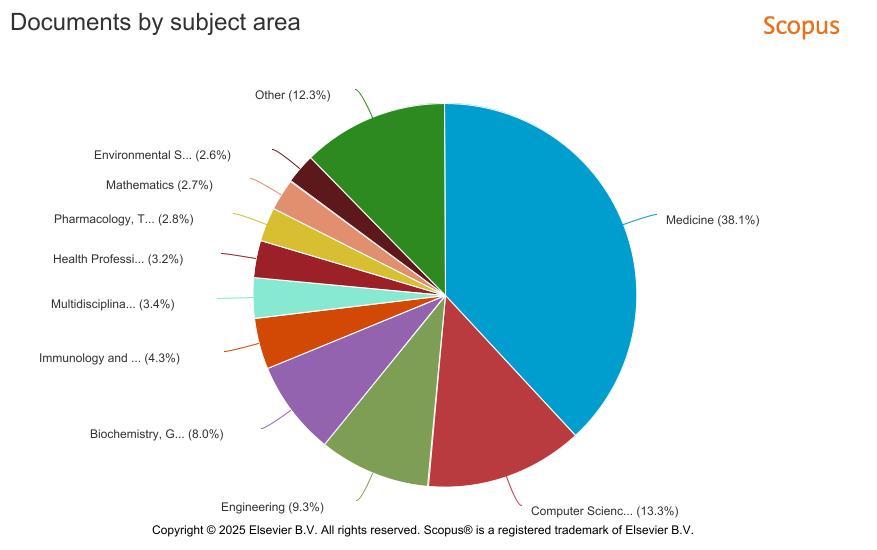

### Scopus analyze by year

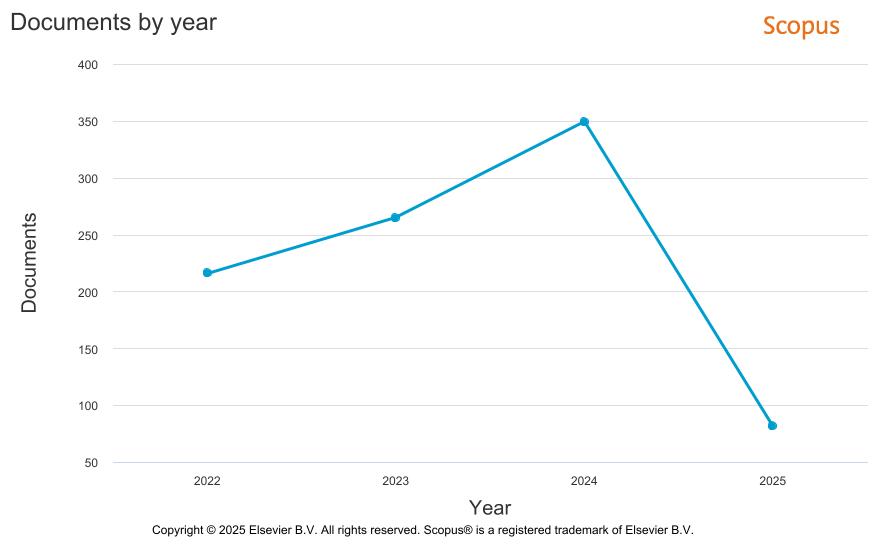

### Scopus analyze by year

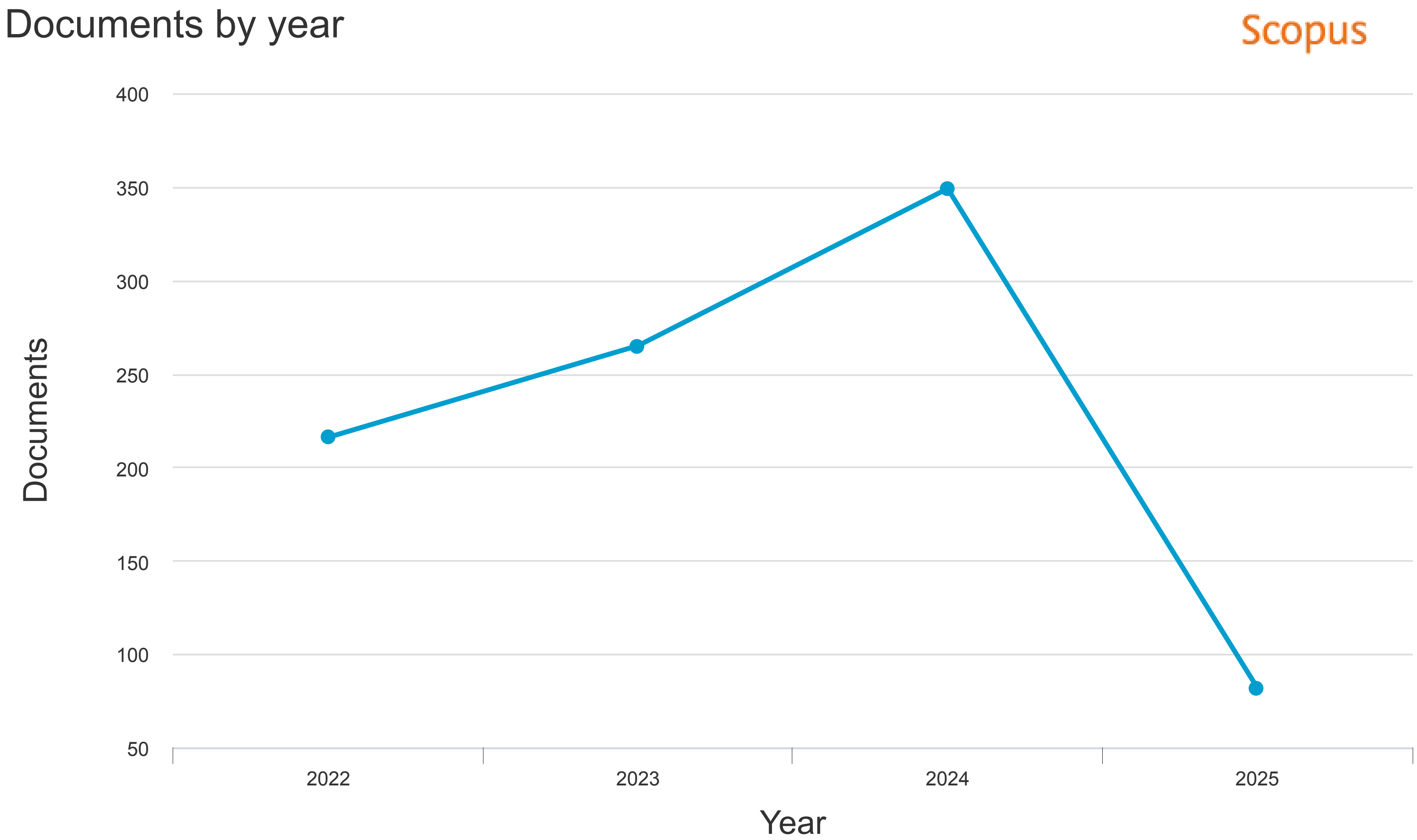


Copyright © 2025 Elsevier B.V. All rights reserved. Scopus® is a registered trademark of Elsevier B.V.
