## Supplementary material for "Revolutionizing COPD and Asthma Management with Artificial Intelligence": Scopus analyze by subject

### Documents by subject area

Scopus

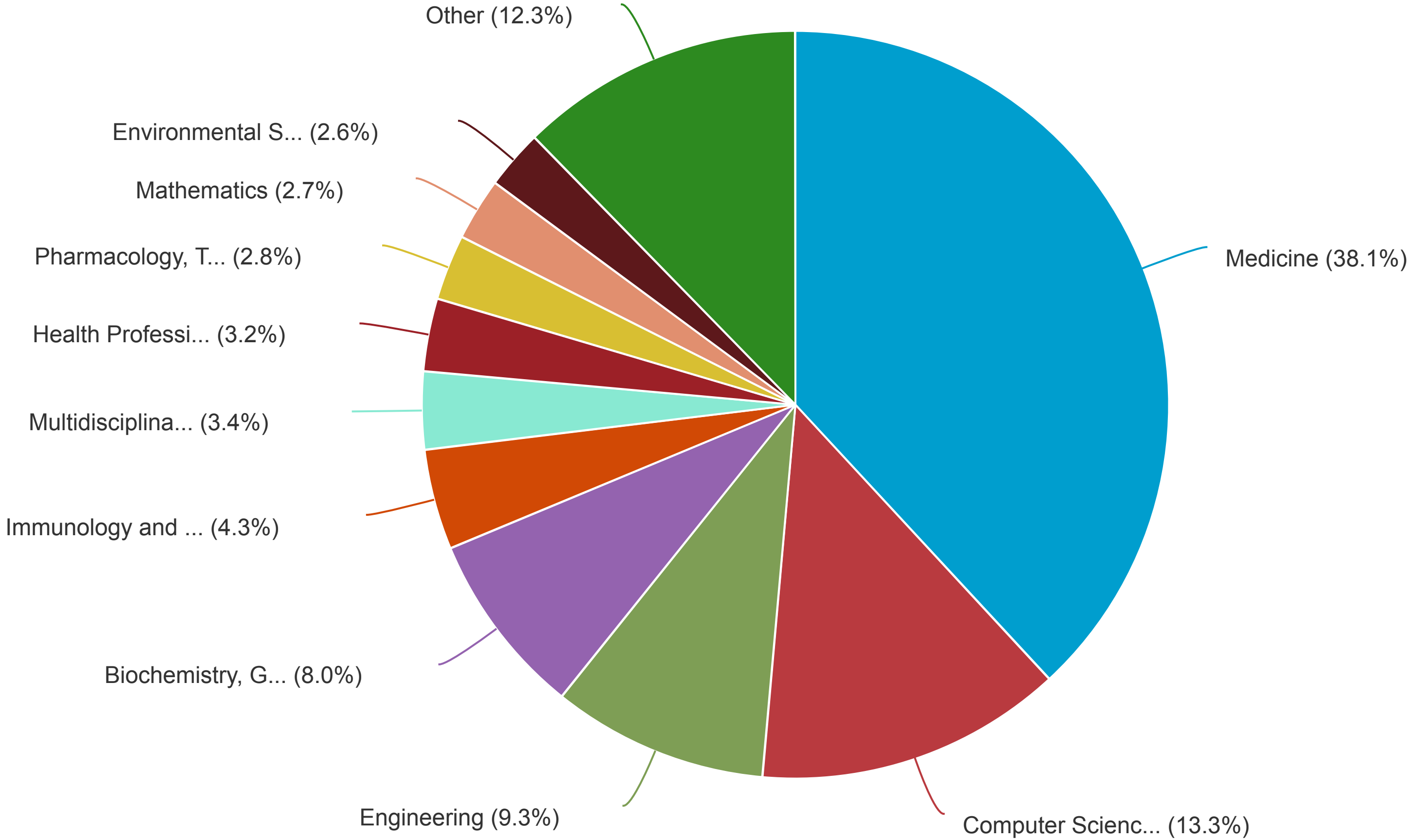


Copyright © 2025 Elsevier B.V. All rights reserved. Scopus® is a registered trademark of Elsevier B.V.
